# Leveraging Foundation Models in Maternal and Child Health: A Systematic Review

**DOI:** 10.1101/2025.08.29.25333768

**Authors:** Xinnie Mai, Yunqian Liu, Philip Chung, Jonathan D. Reiss, Shuang Zhou, Ronald J. Wong, Mingquan Lin, Ivana Maric, Marina Sirota, Nima Aghaeepour, Rui Zhang, David K. Stevenson, Feng Xie

## Abstract

Maternal and child health (MCH) represents a critical domain requiring accurate, timely, and data-driven decision-making to optimize outcomes from pregnancy through early childhood. Foundation models (FMs) are large pre-trained artificial intelligence models that offer potential for clinical support in diagnostics, medical adherence, and reducing disparities. We conducted a systematic review to identify recent studies leveraging FMs in MCH published between 2020 and 2025. Of 785 studies, 63 met the inclusion criteria. FMs demonstrated strong potential to generalize across clinical tasks by integrating multimodal data, including text, electronic health records, imaging, and temporal data to support disease diagnosis, streamline clinical documentation, and generate high-quality medical responses throughout maternal, neonatal, and pediatric care. Moving forward, rigorous validation and close collaboration with clinicians will be essential for the safe, equitable, and effective deployment of FMs in MCH care.

## Introduction

Maternal and child health (MCH) represents a vital healthcare domain characterized by complex, longitudinal care pathways spanning multiple life stages^1–3^, from prenatal care to newborn and pediatric development^1,2^. MCH involves diverse clinical data and requires decision-making that is accurate^4^, timely^4^, and data-driven^4,5^ to improve outcomes for both maternal and pediatric populations. Traditional risk scoring models and machine learning (ML) models such as logistic regression^6^, random forest^7^, and support vector machines (SVM)^8^ as well as deep learning, such as convolutional neural networks (CNN)^9^ and long short-term memory (LSTM)^10,11^, have been applied in MCH to predict outcomes such as preterm birth^12^, low birth weight^13^, gestational diabetes mellitus^7^, anomaly detection^14,15^, and labor complications.^16^ However, these conventional models often rely on hand-crafted features^17^, limited sample size, single data modalities^18^, and struggle with generalizability across diverse populations and institutions^19^.

Recent advances in artificial intelligence (AI)^20^, particularly the emergence of foundation models (FMs), offer transformative potential in overcoming these limitations. FMs are large-scale pretrained models capable of adapting across tasks and domains with minimal fine-tuning. Their capacity for cross-domain generalization, learning transfer, and multimodal data integration makes them particularly well-suited for MCH^21,22^, where data are often sparse, high-dimensional, and distributed over time^22–25^. In MCH, FMs have been applied for transforming care by enabling earlier and more accurate detection of life-threatening conditions, such as pediatric pneumonia^26^ and postpartum post-traumatic stress disorder (PTSD)^27^, through their unique capacity to analyze complex data. This enhanced analytical power ultimately contributes to improved survival and reduced long-term complications for both mothers and infants.

Representative FM architectures include Generative Pre-trained Transformer (GPT)^28^, Bidirectional Encoder Representations from Transformers (BERT)^29,30^, Vision Transformer (ViT)^31,32^ and Large Language Model Meta AI (LLaMA)^33,34^. These models have demonstrated improved performance in information retrieval^35^, disease diagnosis^31,32,36,37,38^, clinical documentation^39^, and generation of patient-friendly educational content ^40–42^. Their scalability, data processing capability^43–45^ and adaptability make them well-suited for the longitudinal and multimodal nature of MCH, offering a unique opportunity to unify fragmented clinical information and support more holistic, real-time decision making. By bridging data silos, integrating diverse information sources, and enabling timely, personalized interventions, these models hold significant potential to enhance maternal and neonatal health outcomes. Rather than replacing existing tools, these models can augment clinical workflows by extracting actionable insights from complex, multimodal datasets across prenatal, neonatal, and pediatric stages.^146^

While several recent reviews have assessed the use of FMs in general medicine and healthcare domains^47,23,48^, most MCH focused reviews to date have centered on traditional ML approaches.^49,50,17^ These conventional models typically address narrow, task-specific problems, and lack of the generalizability, multimodal capacity, and contextual reasoning that FMs can offer. Despite the growing interest in FMs, a lack of comprehensive evaluation of their potentials to improve the outcomes across the maternal and pediatric care continuum remains.^5,51^ This review addresses this gap by consolidating the latest advancements in FM applications for MCH^1,3,52^, systematically evaluating their performance, limitations, and future directions for safe and equitable implementation in real-world clinical care.^52,53^ We outline persistent gaps in multimodal integration^1,54^, real-world validation^55,56^, and ethical considerations.^27,52,57,58^ Finally, we propose strategic directions for future research to optimize the implementation of FMs in MCH, emphasizing the importance of domain-specific fine-tuning, multimodal learning, and robust clinical validation.^26,27,54,55^

## Methods

### Search Strategy and Data Sources

We conducted a systematic review by including all studies applying FMs to MCH, and identified studies from four databases: PubMed, Web of Science, Embase, and Scopus. Our literature search covered publications from January 1, 2020, to January 31, 2025. The search strategy was developed following the Preferred Reporting Items for Systematic Reviews and Meta-Analyses (PRISMA) guidelines^59^, with the full search term listed in the Supplementary Materials. This systematic review was registered with PROSPERO (CRD 1104501).

### Inclusion and Exclusion Criteria

Studies were included if they focused on applying FMs or large-scale pre-trained models in maternal, perinatal, neonatal, pediatric, or child health settings. Eligible original studies presented clearly defined model architectures, data sources, and evaluations. We included only studies published in English with full-text availability. Studies were excluded if: (1) unrelated to maternal or child health; (2) did not involve FMs; (3) were non-primary research publications (e.g., reviews, editorials, or conference abstracts); (4) were not published in English; or (5) had no access to available full text. Two reviewers (XM and YL) conducted the screening process independently and assessed titles and abstracts before conducting full-text evaluations to determine eligibility. Any disagreements between reviewers were resolved through discussion or, when necessary, by consulting a third reviewer with expertise in health informatics (FX) to reach a consensus.

### Data Extraction and Analysis

After full-text screening, relevant data were systematically extracted. First, we extracted the name of the FMs used in each study (e.g., BERT, GPT, or LLaMA). Second, regarding data characteristics, we recorded the input data modality (e.g., text-based data, structured electronic health records [EHRs], clinical notes, social media content, and multimodal data), the sample size used for fine-tuning, the clinical application described in the article (e.g., diagnosis support, adherence monitoring), and its corresponding clinical application category (such as clinical natural language process [NLP] or decision support). Furthermore, we also recorded the clinical task of each article, including text extraction, medical image classification, and clinical evaluation. We also captured the relevant medical or disease branch, whether the study was directly related to maternal or child health. Lastly, the extracted data were systematically compiled to facilitate a comprehensive analysis. To facilitate a structured synthesis, studies were grouped into three overarching application domains based on their primary objective: (1) clinical NLP; (2) medical question answering and knowledge assessment; and (3) predictive modeling. Clinical NLP encompasses tasks involving information retrieval, text summarization, and text classification. Medical question answering and knowledge assessment involve using FMs to support diagnostic reasoning, guideline adherence, and patient-specific decision-making. Predictive modeling focuses on applying FMs to predict outcomes or diagnose disease by analyzing complex medical data, including clinical text, imaging, structured EHRs, temporal data and multimodal data.

## Results

### Selection Process and Results Overview

Our initial search yielded 1,164 articles, of which 379 duplicates were removed, leaving 785 articles for title and abstract screening. During this phase, 717 articles were excluded for the following reasons: not related to MCH (n = 339), not involving FMs (n = 340), incorrect publication type (n = 35), or not published in English (n = 3). This resulted in 68 full-text articles being assessed for eligibility. Among these, 5 articles were excluded due to incorrect publication type (n = 2), no access to full text (n = 2) or lack of relevance to FMs (n = 1). Ultimately, 63 articles met the inclusion criteria and were included in the final analysis (Table 1). The PRISMA flow diagram detailing the study selection process is presented in Figure 1. Analysis of the publication trends revealed a steady increase in research activity over recent years, with 36 of the 63 included studies published in 2024 alone. This had reflected a growing research interest in applying FMs to MCH (Figure 2a).

**Figure 1:**
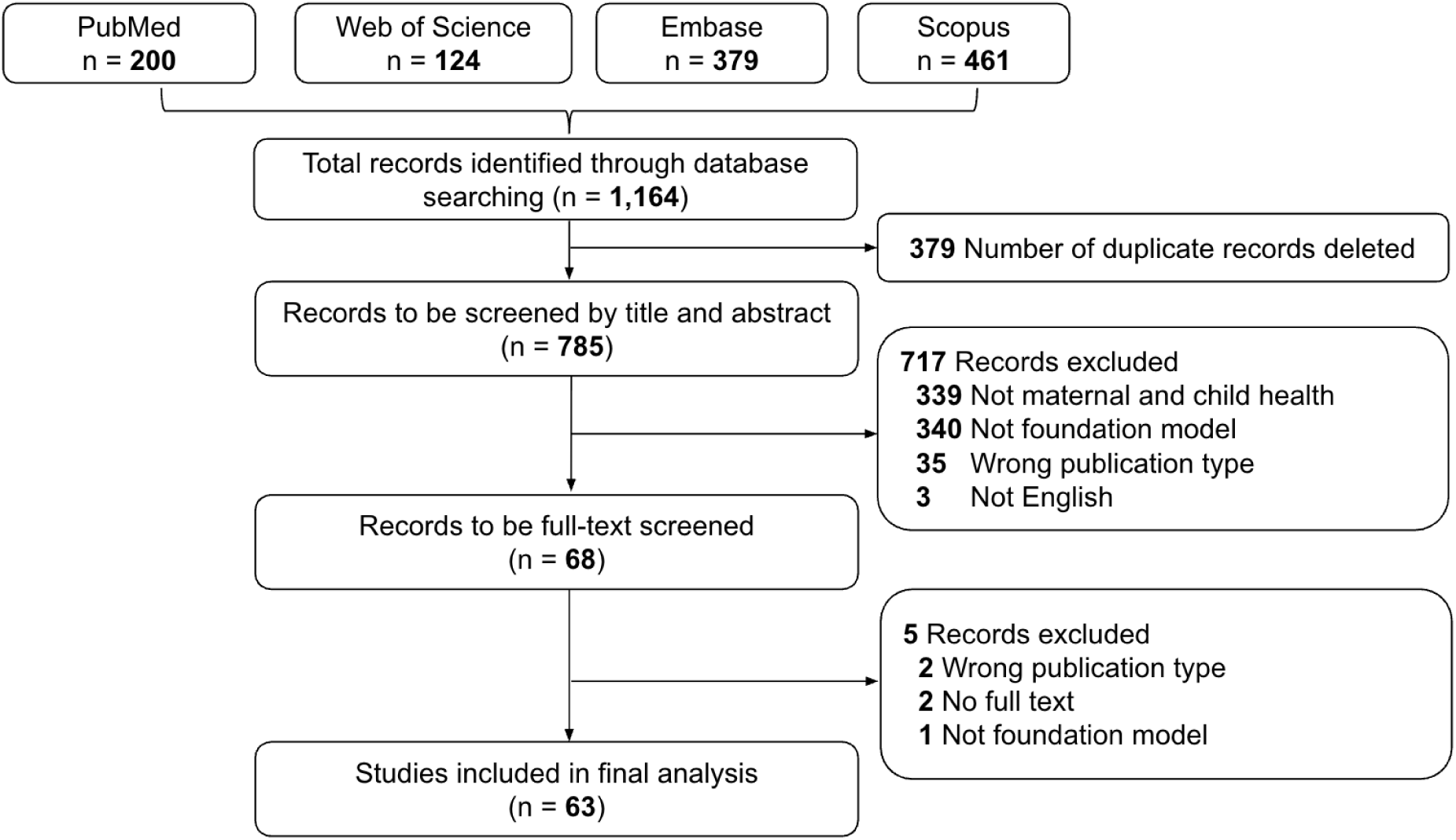
PRISMA Flow Diagram of Study Selection and Screening Process. PRISMA flowchart showing the study selection process.

**Figure 2:**
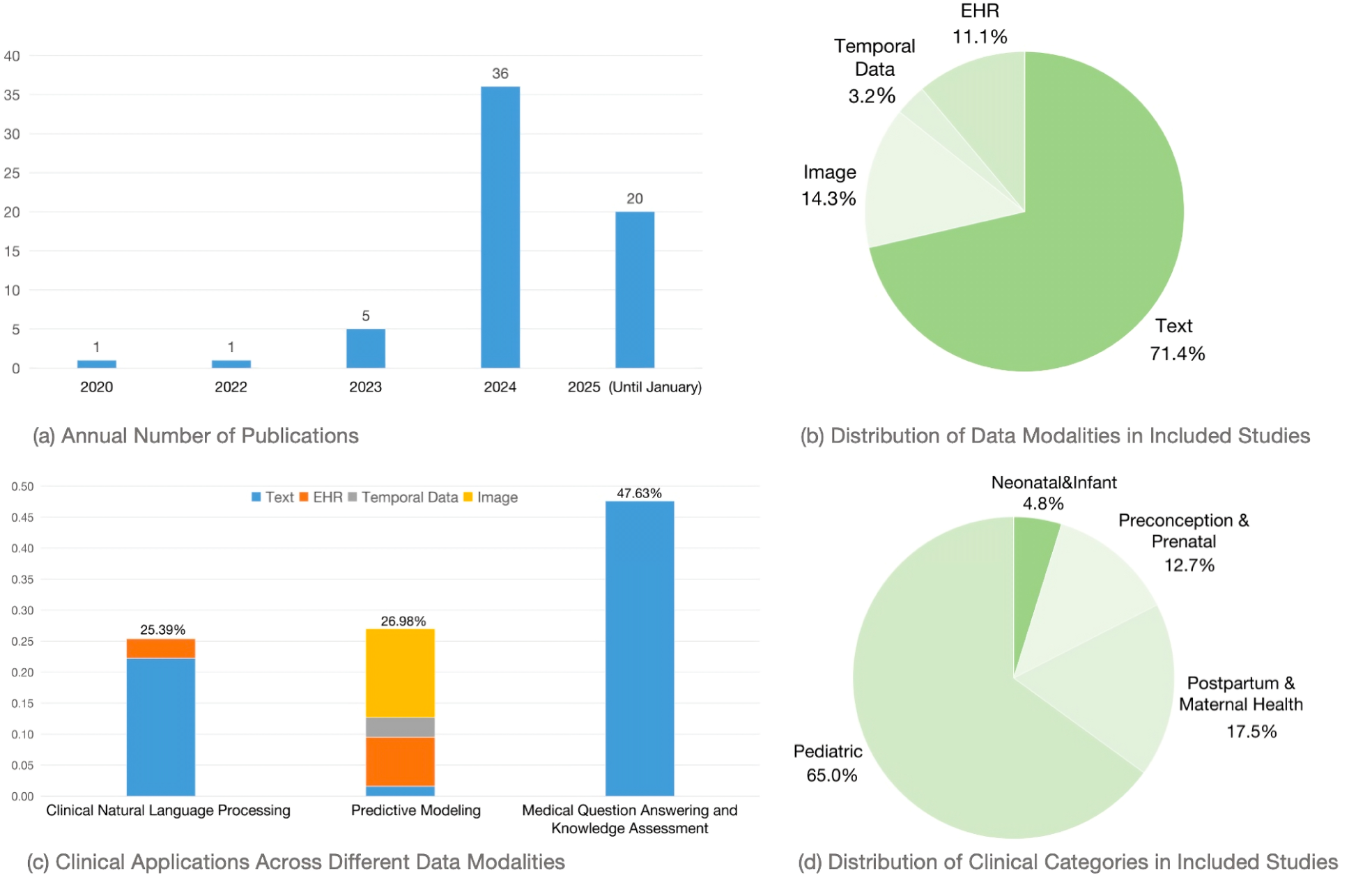
Applications and Trends of Foundation Models in Maternal and Child Health Across Clinical Use Cases and Data Modality. This figure summarizes key patterns in the included studies. (a) Annual number of publications on foundation models in maternal and child health. (b) Distribution of data modalities in included studies. (c) Summary of clinical applications across modality. (d) Distribution of clinical categories in included studies.

**Table 1:**
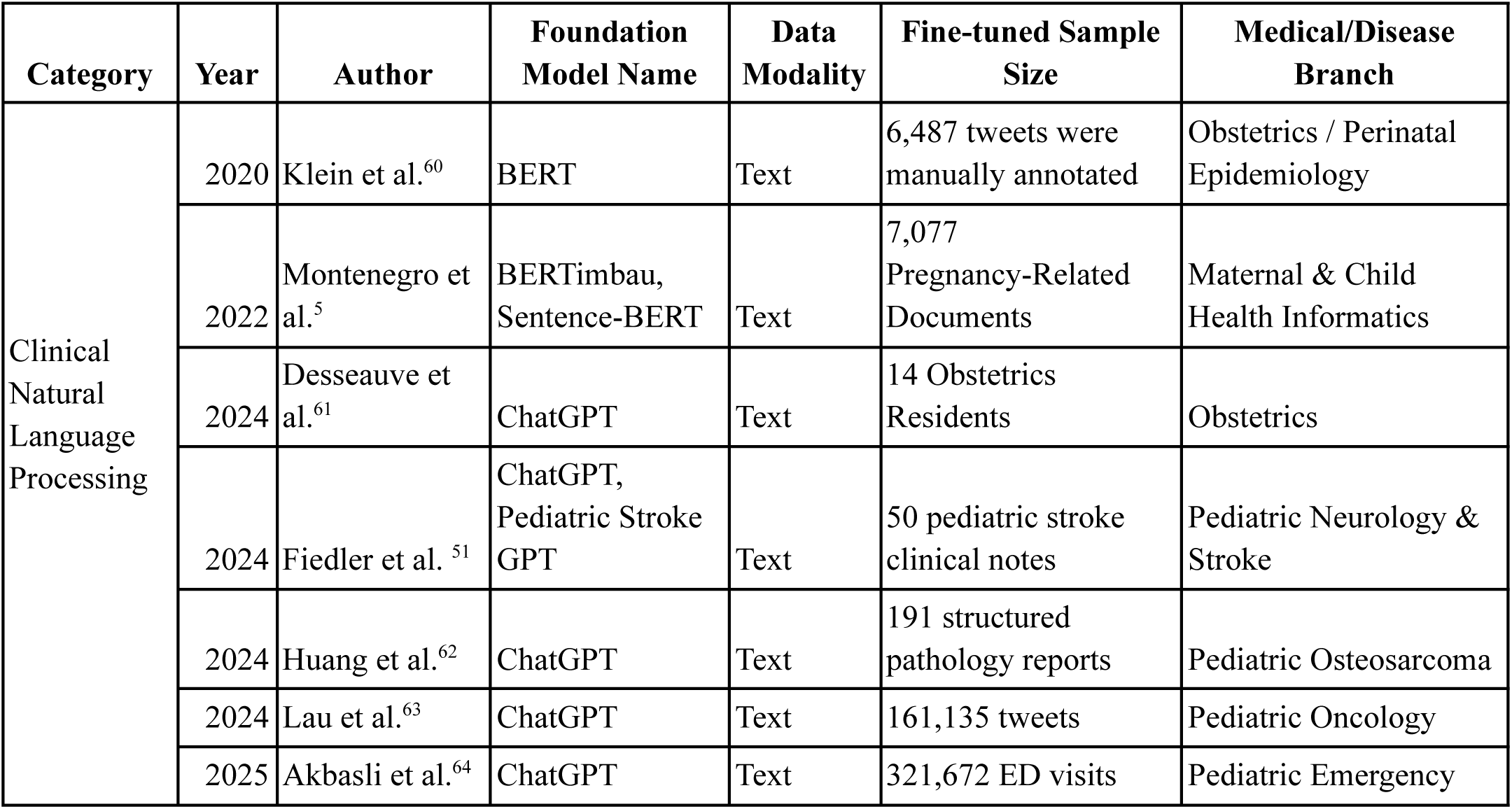
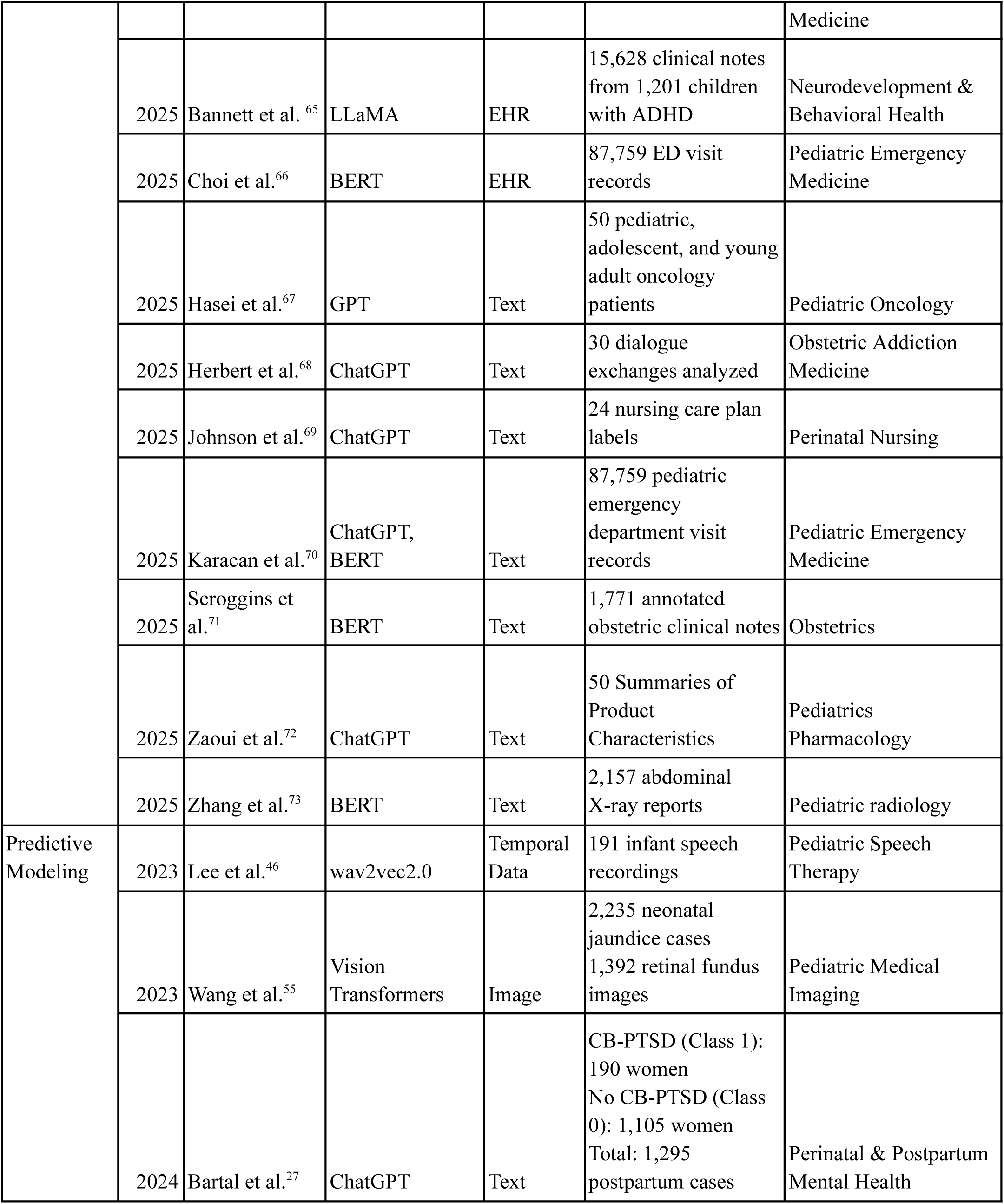
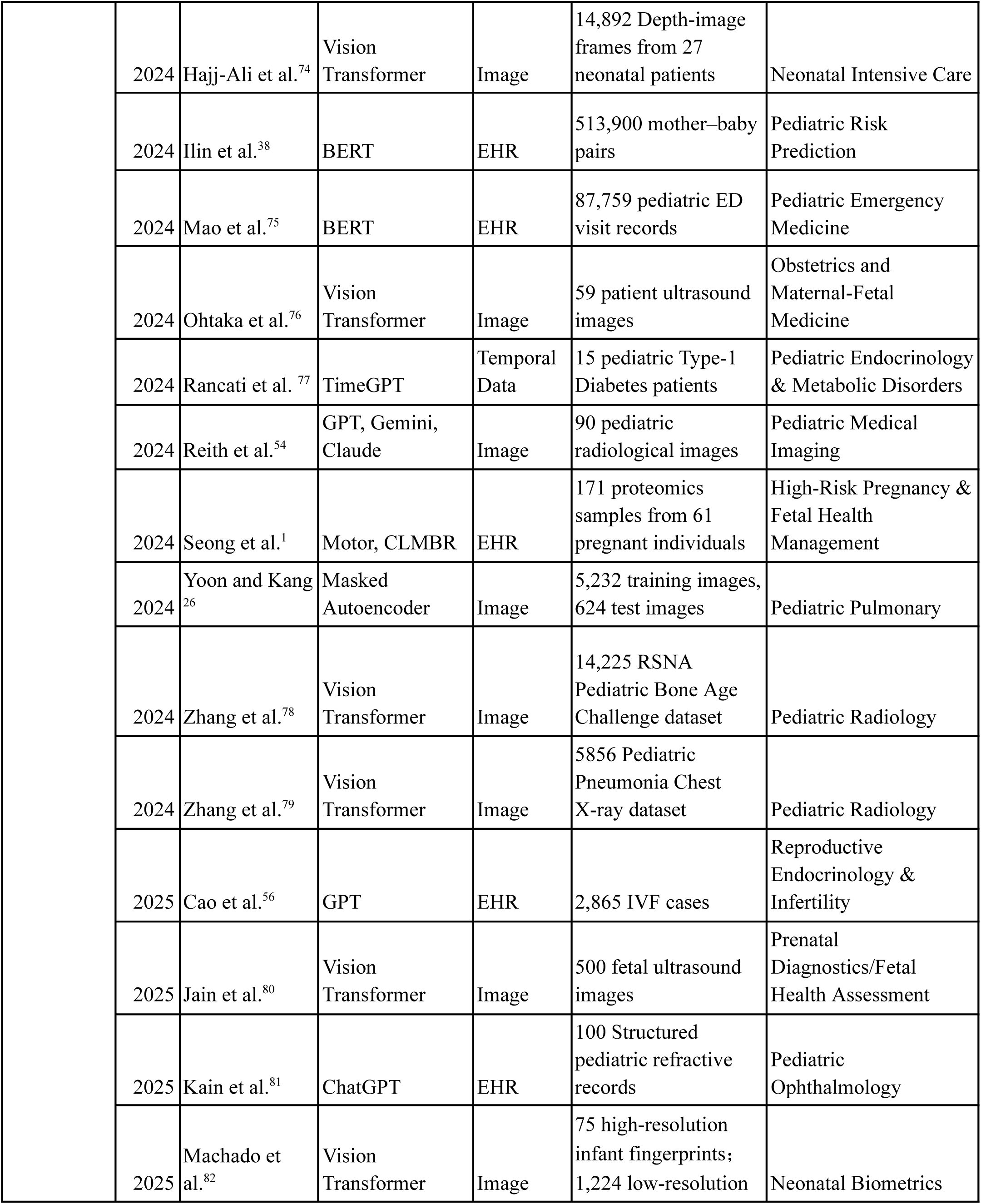
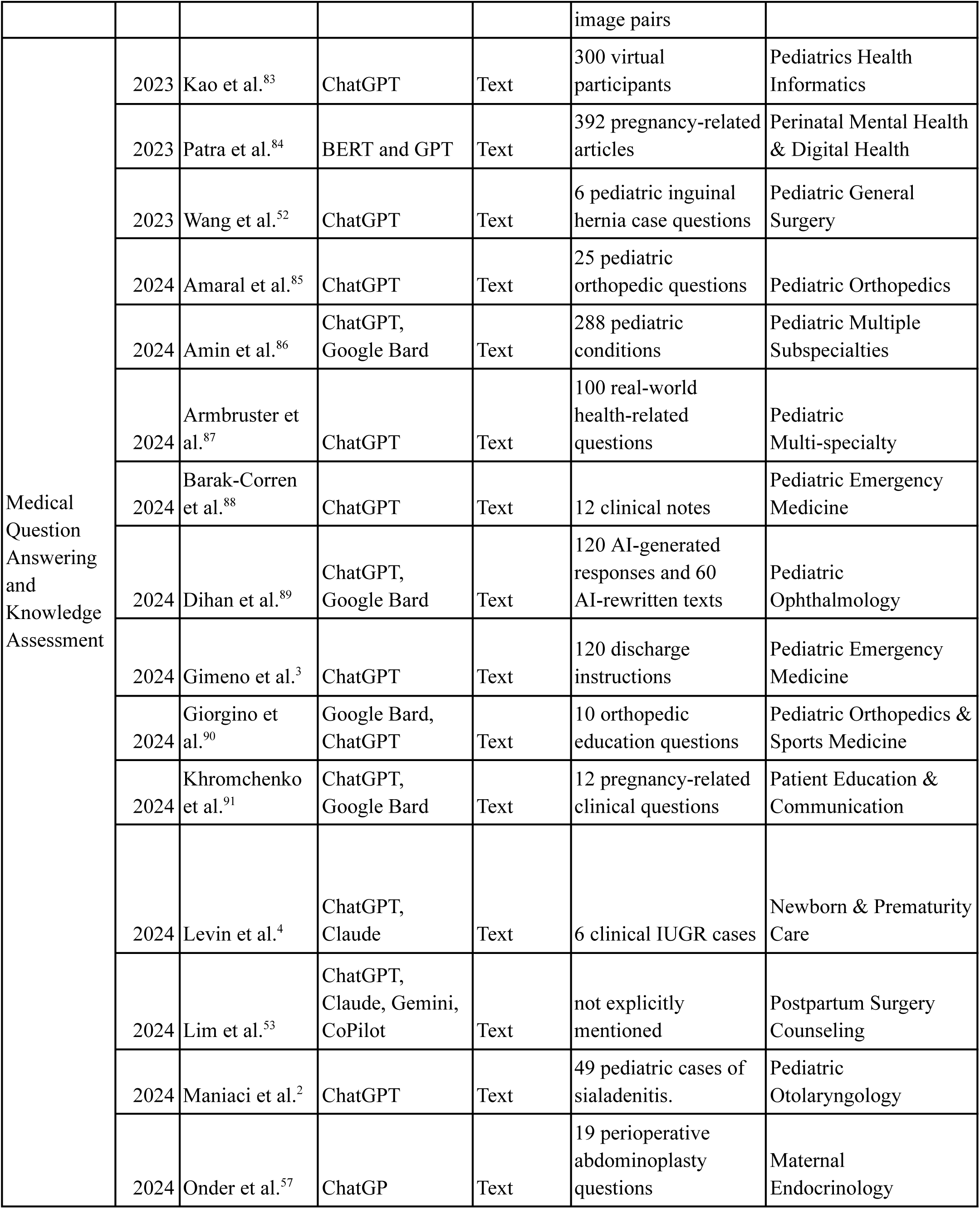
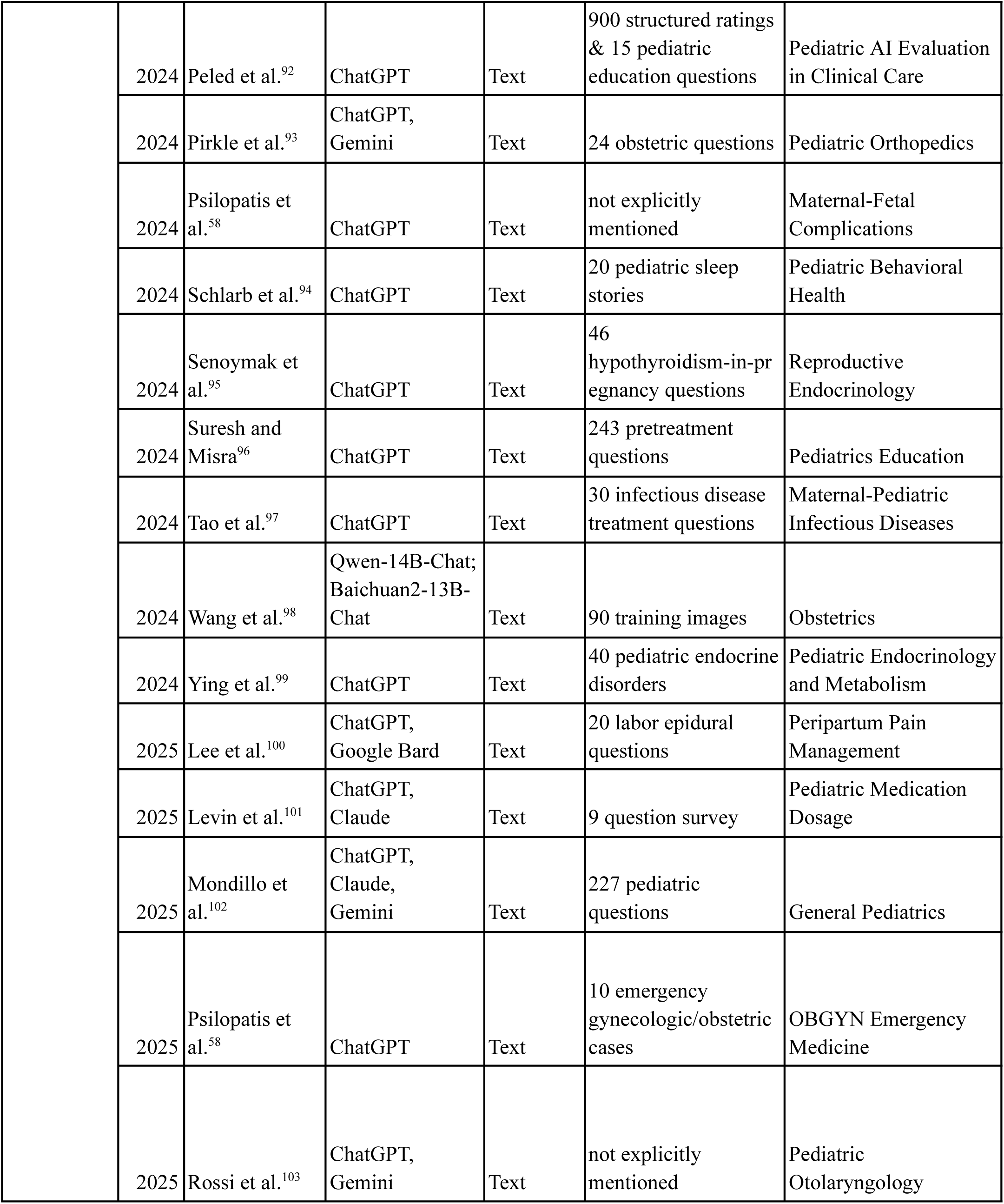
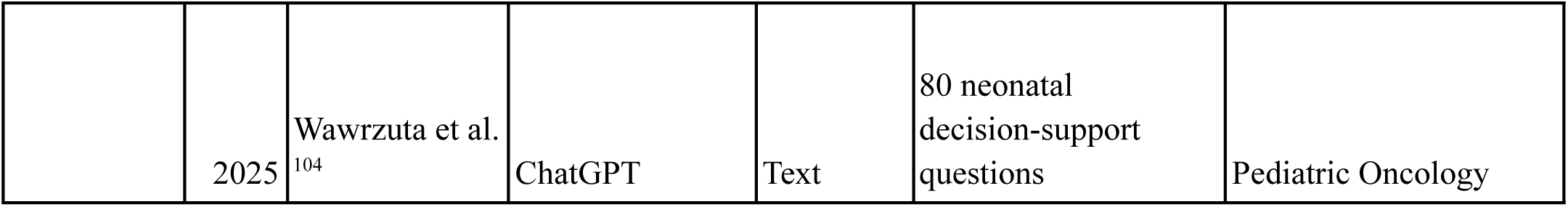
Summary of Included Studies on Foundation Models Applied to Maternal and Child Health.

FMs are large-scale deep learning models that use transformer-based architectures to learn from large amounts of unlabeled data, which can be fine-tuned for specific clinical tasks. In MCH, FMs support diagnostic prediction, clinical text analysis, and patient monitoring across diverse data types. Table 2 summarizes key FMs used in MCH, including GPT^28,105^, BERT^29^, LLaMA^33^, clinical language model based representations (CLMBR)^1,106^, ViT^31,55^, and wav2vec 2.0^46,107^. These models differ in mechanism^29^ and are therefore suited to distinct task domains across text, image, and speech modalities. For example, GPT and LLaMA use a decoder-only transformer^28,105,33^, BERT uses bidirectional encoders, and ViT applies transformers to image patches^31^. These applications span summarizing pediatric stroke notes, extracting osteosarcoma reports, analyzing Attention-Deficit/Hyperactivity Disorder (ADHD) medication adherence, generating pregnancy-related clinical timelines from EHR, identifying maternal health topics, and detecting autism from children’s speech.

**Table 2:**
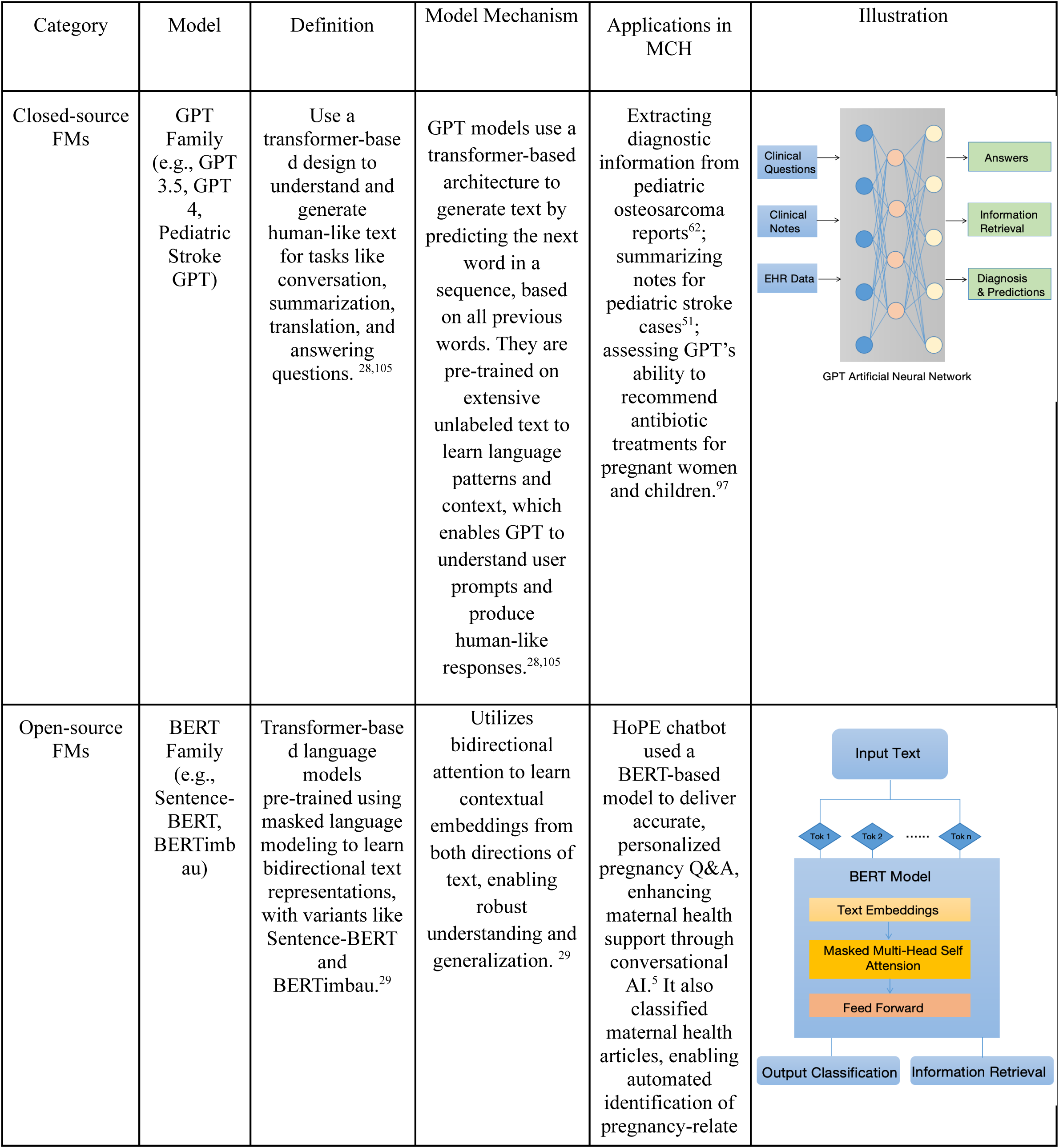
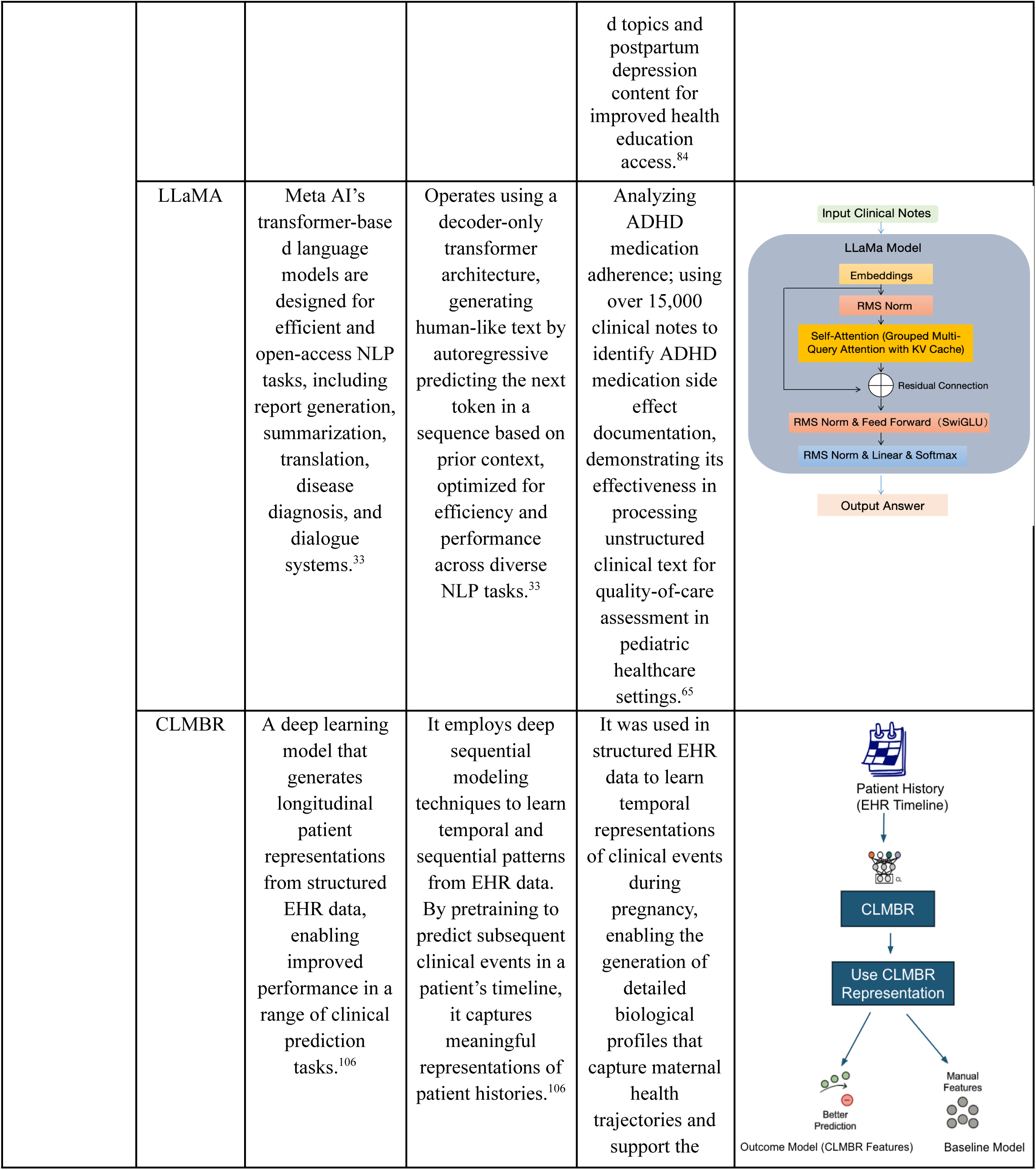
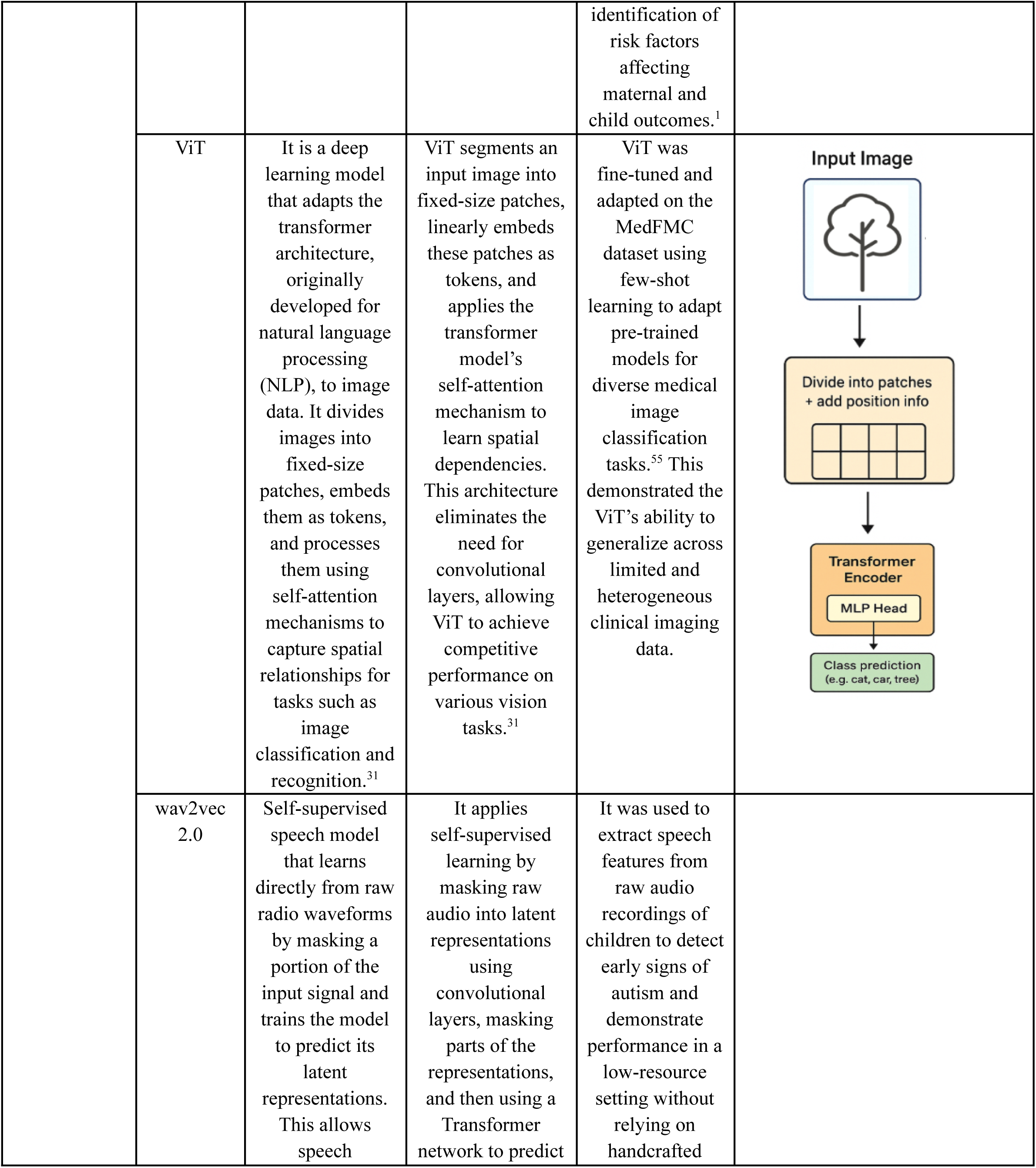
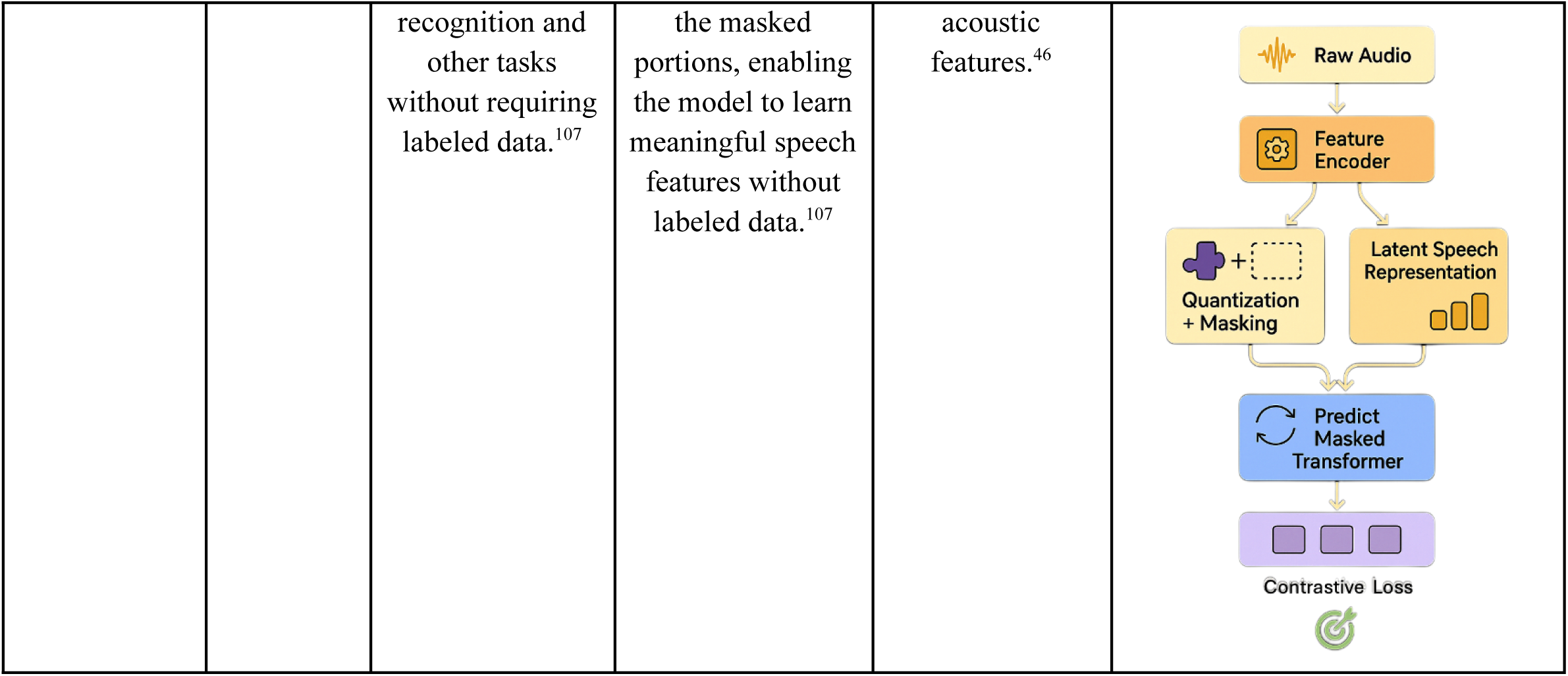
Overview of Foundation Models in Maternal and Child Health.

### Analysis of Data Modalities of the Foundation Models

FMs have emerged as a growing focus in MCH research, with notable acceleration in publication activity in 2024 (Figure 2a). Our analysis revealed that 71.4% (45/63) of reviewed studies utilized text-based FMs, primarily from the BERT^5,84^, GPT^52,83,108^, and LLaMA^65^ families, which are widely adopted for processing unstructured clinical data and supporting clinical decision-making (Figure 2b). GPT-3.5 and GPT-4 variants, for instance, achieved over 98% accuracy in extracting clinical notes from pediatric osteosarcoma reports^62^. Fine-tuned models like Pediatric Stroke GPT^51^ have been applied to summarizing neurological findings, while Sentence-BERT^5^ has enabled applications in maternal health education and multimodal classification. LLaMA^65^ has also been employed in pediatric behavioral health to assess medication adherence and improve documentation quality.

Visual FMs are also increasingly used (14.3%, 9/63) in MCH imaging tasks (Figure 2b). ViTs have shown strong performance in pediatric imaging tasks, particularly in few-shot and zero-shot learning scenarios where annotated data is limited.^55^ Similarly, Masked Autoencoders (MAEs), pretrained on adult chest X-rays and fine-tuned on pediatric datasets, achieved high diagnostic accuracy in pneumonia detection (area under the curve [AUC] = 0.996). Vision-language models are also emerging to enhance diagnostic interpretability by linking imaging with textual findings.^55,76,79^

Beyond text and imaging, a smaller but growing number of studies have explored additional modalities such as structured EHRs (11.1%, 7/63) and temporal data (3.2%, 2/63). Models like Time-to-event Risk (MOTOR)^1^ and CLMBR^1^ utilize longitudinal EHR data to predict maternal blood protein levels, enabling dynamic pregnancy risk stratification. Speech-based FMs, such as wav2vec 2.0^46^, have been applied to detect autism spectrum disorder by analyzing child vocal features. In addition, emerging multimodal systems, such as Gemini^53,54,93^, Bard^89,91,100^, and Copilot^53^, integrate textual, visual, and contextual inputs to support clinical reasoning, diagnostic decision-making, and patient education. These trends reflect growing interest in multimodal FMs while highlighting underexplored opportunities beyond text-dominant applications.

### Analysis of Clinical Applications in MCH

FMs increasingly support various clinical applications in MCH, enabling more efficient, personalized, and equitable care. As illustrated in Figure 2c, the majority of studies (47.63%, 30/63) focused on medical question answering and knowledge assessment, followed by predictive modeling (26.98%, 17/63) and clinical NLP (25.39%, 16/63). These models were used to generate medical recommendations^26,84,92^, assist in decision-making^27,56,58^, and enhance communication^5,84,95^ demonstrating the strengths of FMs in language interpretation^5,84,95^, summarization^5,26,58^, and real-time generative response. As shown in Figure 2d, among the 63 studies reviewed, 65.0% (41/63) focused on pediatric care, while 4.8% (3/63) examined neonatal and infant health. Postpartum and maternal health was covered in 17.5% (11/63) of studies, and 12.7% (8/63) addressed preconception or prenatal care. This distribution indicates that pediatric research is currently the most explored area within the MCH domain, with fewer studies focusing on earlier life stages. To synthesize the diverse applications of FMs across maternal, newborn, and pediatric care, Figure 3 visually maps commonly used FMs, such as GPT, BERT, LLaMA, ViT, CLMBR, and wav2vec 2.0, to represent the clinical use cases across different clinical tasks, including prenatal risk prediction, neonatal intensive care units (NICU) triage, and pediatric medication guidance.

**Figure 3:**
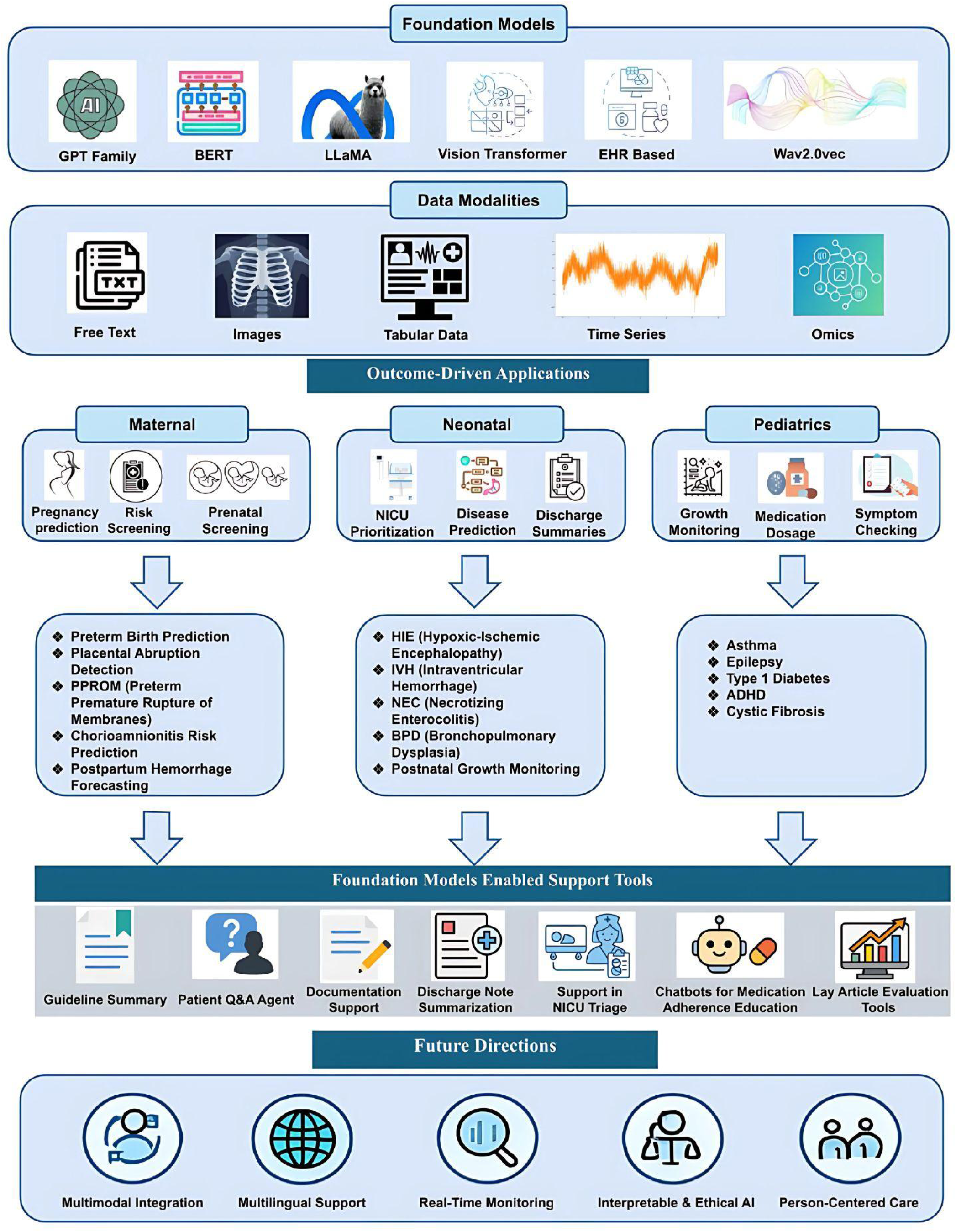
Applications of Foundation Models in Maternal and Child Health. This figure illustrates representative FMs architectures, supported data modalities, enabled support tools, outcome-driven applications across maternal, neonatal, and pediatric care, and key future directions including multimodal integration and ethical, interpretable AI.

In maternal care, FMs have been increasingly adopted to support earlier risk detection and individualized prenatal care^109^. Applied to structured EHR data, unstructured clinical notes, and patient-reported information, these models can predict pregnancy complications ^5,56^, monitor high-risk pregnancies ^27,58,92^, and provide tailored responses to medical questions related to MCH^5,84,95^. One representative example is the High-Optimization and Precision Medicine (HoPE) conversational agent, a real-time mobile chatbot designed to support prenatal care. It combines BERTimbau and Sentence-BERT in a dual-encoder architecture to retrieve personalized, evidence-based responses from a curated corpus of over 7,000 medical documents related to pregnancy^5^. Early evaluations reported high alignment between HoPE’s and healthcare professionals’ recommendations, highlighting its potential for trustworthy at-home counseling, which is particularly useful in low-resource or rural settings, where access to clinical expertise may be limited. Beyond HoPE, FMs such as GPT and BERT have also been applied to interpret and summarize obstetric guidelines, answer patient questions about pregnancy complications and prenatal care, and evaluate the quality of lay health articles on reproductive health topics. These tools reduce documentation burden, deliver individualized support, and contribute to more informed, equitable, and efficient reproductive healthcare.

In newborn care, FMs enhance clinical decision-making by analyzing labor, delivery, and the early neonatal period data to detect complications and guide timely interventions.^1,2,46,55,63^ In one study, a FM, ViT was used to classify over 2,200 pediatric and neonatal medical images, including those related to neonatal jaundice^55^, demonstrating strong performance in identifying skin-related conditions from imaging data. Beyond image classification, multimodal FMs show considerable potential in integrating EHRs^1,65^, physiological signals^88^, and imaging data to inform complex perinatal decisions.^26,54,55^ Their use during and immediately after delivery facilitates early risk detection and supports a seamless transition in care from the maternal to the neonatal stage.

In pediatrics, FMs have been deployed across various specialties to assist in diagnostics, monitor treatment adherence, and streamline clinical documentation.^110^ These models have proven effective in classifying developmental^46,65^ and neurological disorders^51,54^, extracting structured variables from unstructured clinical notes^62,92^, and personalizing care plans for chronic conditions such as ADHD.^65^ In oncology, LLMs, such as GPT-4, have been used to extract diagnostic information from pathology reports in pediatric osteosarcoma.^62^ Concurrently, domain-specific LLMs, such as Pediatric Stroke GPT, have analyzed clinical narratives to assist in neurologic assessments for pediatric stroke.^51^ Applications in behavioral health include leveraging models, like LLaMA to evaluate medication adherence in children with ADHD, thereby enhancing clinician feedback and individualized tracking.^65^ FMs have also contributed to pediatric oncology and health communication by synthesizing large volumes of unstructured data, including summarizing and classifying social media content related to childhood cancer. ^65,104,3,63^

## Discussion

The rapid evolution of FMs has generated significant interest in their application across MCH, a field defined by complex, longitudinal, and high-stakes care trajectories^111^. These models offer promising opportunities to enhance clinical decision-making, reduce documentation burdens, and support personalized interventions.^112^ This review highlights the current dominance of language-based FMs in MCH, alongside emerging efforts to incorporate multimodal inputs such as imaging, temporal, and structured health data. While clinical evaluation and risk prediction are longstanding priorities in MCH, FMs integrate multimodal data, capturing nuanced trends over time, and generate scalable insights across care settings. This systematic review synthesizes the current landscape of FM applications in MCH, underscoring their transformative potential and the challenges that must be addressed to ensure safe and equitable clinical integration.

A key strength of FMs lies in their ability to integrate diverse data modalities, including unstructured clinical text^5^, structured EHRs^51,113^, and medical imaging such as chest X-rays.^26^ This capability is particularly relevant in MCH, where effective decision-making requires the synthesis of diverse and longitudinal information, such as prenatal ultrasound images, structured EHRs, developmental assessments, and physician notes. FMs have demonstrated strong potential to move beyond text-only inputs by integrating underutilized modalities. Recent models have shown encouraging progress in combining these heterogeneous inputs^1^ to support complex prenatal and pediatric diagnostics. Continued progress in this area offers the opportunity to develop models that learn shared cross-modal representations, unlocking the greater adaptability and reliability in diverse MCH settings.^114^ Multimodal FMs can also help move the field toward “world models”^115^ for MCH, systems capable of understanding and reasoning across the full patient context, integrating temporal, multimodal, and cross-domain information.^116^

Furthermore, FMs perform effectively with limited labeled data^117^, a notable advantage in MCH, where expert annotations are often challenging to obtain. This scarcity stems from the relative rarity of certain maternal or pediatric conditions, stringent privacy regulations, and ethical challenges in collecting data during pregnancy or early childhood^118^. By leveraging large-scale pretraining and efficient adaptation, FMs can address these limitations and generalize to downstream tasks with minimal supervision.^26^ For example, few-shot learning approaches such as Meta-Baseline and Visual Prompt Tuning^55^ have shown strong performance in neonatal jaundice classification using only 1, 5, or 10 labeled samples per class^55^. Expanding the application of low supervision methods to additional MCH-relevant tasks represents a promising and underexplored direction for future research. This includes areas, such as fetal imaging, which demands time-intensive expert annotation; development surveillance, which depends on long-term tracking; and postpartum risk prediction, where outcome data can be infrequent, delayed, or inconsistently documented.^119,120^

FMs contribute to improving workflow efficiency in MCH by automating documentation tasks such as delivery summaries, pediatric discharge notes, postpartum progress reports, and translating clinical jargon into patient friendly language. In parallel, models like GPT, Bard, Gemini, Claude, and Copilot have been increasingly evaluated for their ability to generate accurate, accessible responses to medical questions, which supports patient education and clinical knowledge assessment across pregnancy^57^, labor^100^, postpartum care^84^, and pediatric conditions.^4,52^ These capabilities are crucial in high-pressure environments, such as NICU and postpartum care, where continuity and clarity of information are paramount. FMs support clinical documentation by condensing complex medical content into structured, comprehensible summaries, enabling clinicians to manage workload while maintaining patient-centered communication.^51,62^ By simplifying complex clinical information, FMs can enhance a patient’s ability to engage with their care and make informed choices. The ability of FMs to generate multilingual and culturally sensitive content further enhances their role in promoting health equity.^121^ By bridging communication gaps across diverse patient populations, these models can help ensure that care instructions and educational materials are accessible to users with varying levels of health literacy.^84,95^ In MCH, one emerging strength of FMs is their ability to generate context aware responses that adapt to patient’s stage of care, such as pregnancy trimesters, postpartum phases, or child development milestones. This follows FM, like conversational tools and patient portals to deliver timely and relevant answers that complement with the clinical encounters and reinforce the continuity of care.^5,84^ These capabilities illustrate the growing role of FMs in supporting medical question answering and patient communication within maternal and pediatric care.

FMs enhance clinical decision support in MCH by interpreting clinical notes, suggesting diagnoses, and assisting with triage and treatment planning. FMs have been applied across MCH contexts, such as pediatric stroke diagnosis^51^, ADHD documentation review^65^, and maternal-fetal risk assessment to reduce clinician cognitive burden, improve diagnostic accuracy^26,54^, and provide real-time decision support by identifying high-risk patients and generating triage recommendations from incomplete data inputs^58,92^. These capabilities are particularly valuable in time-sensitive, resource-constrained settings, where scalable clinical reasoning and adherence to evidence-based guidelines are critical. While FM applications in MCH remain nascent, LLMs like GPT-4 have demonstrated superior diagnostic performance to physicians in adult emergency and internal medicine^122–124^. Multimodal FMs have also shown promise in integrating clinical notes, structured data, and imaging for triage and diagnostic reasoning in adult care, offering a compelling foundation for their future adaptation in MCH workflows.

Existing clinical decision support tools offer useful analogies for FM integration. For example, the BiliTool algorithm^125,126^ is widely embedded in pediatric practice to assess neonatal jaundice risk and guide phototherapy decisions based on structured, guideline-based thresholds^126,127^. Emerging FM-based approaches extend this paradigm by automating skin-image-based jaundice assessments using ViTs, enhancing triage accuracy and workflow efficiency^55^. Similarly, antenatal corticosteroid algorithms assist clinicians in balancing the risks and benefits of preterm steroid administration^128^. When embedded in EHRs, these tools align care with medical guidelines. FMs trained on longitudinal health data, such as CLMBR and MOTOR could further personalize such decisions by identifying individualized risk patterns and generating explainable treatment recommendations. Future clinical decision support FM-based systems in MCH may adopt similar integration models, delivering transparent outputs that clinicians can verify and adjust in real time^129–131^. FMs have shown promise in optimizing complex clinical decisions, such as personalized total parenteral nutrition (TPN) planning in NICUs^132^, by integrating multivariate patient data. They also enable multimodal data integration across EHRs and omics, enhancing molecular association studies through linked clinical and immunological data^133^. Additionally, FMs support real-time physiological monitoring using wearable data in pregnancy, identifying patterns linked to prematurity risk and enabling early warning systems in maternal health^134^. Embedding these tools into EHRs and clinician-facing interfaces, along with robust validation and user training, will be essential to ensure FMs function as reliable and trustworthy collaborators in care delivery.

### Limitations and Challenges

Despite their potential, several limitations of FMs remain in MCH. An area that merits further attention when applying FMs to MCH is the clarity and contextual adequacy.^92^ While LLMs often generate fluent responses^53^, they may occasionally omit key clinical details, rely on unsupported references, or use overly technical language^52,95^, which highlight the need for external validation^135,136^, particularly for patient-facing or clinical communication. Validating model performance across real-world contexts and diverse populations can help ensure that FMs enhance, rather than hinder, clinical care. Additionally, the black-box nature of many FMs may pose challenges to building clinical trust. Clinicians remain cautious without transparent justifications for recommendations^137,138^, especially in high-stakes scenarios where contextual reasoning is essential.^92,95^ To facilitate responsible clinical adoption, models should be utilized under expert supervision and integrated into systems that ensure clinical validity, explainability, and fairness.^53,58,88^ Techniques such as evidence citation, confidence scoring, and interpretable architectures are necessary to achieve this goal.^139–141^ Explanations must be adapted to different audiences: simplified and multilingual for patients, and technically detailed for providers.

Moreover, many commonly used FMs have not been fine-tuned^24,142,143,144^ on domain-specific data related to maternal-fetal physiology or pediatric care, which limits their reliability when applied to conditions such as gestational diabetes, intrauterine growth restriction, or pediatric ADHD.^65,145,146^ These limitations are particularly critical in neonatology, where diseases are often rare, developmentally dynamic, and physiologically distinct from MCH populations. Without targeted training or fine-tuning on MCH specific datasets, these general domain models may encounter shifts and fail to generalize accurately. Therefore, this leads to degraded performance in real-world applications. Despite promising results in general medical tasks, there remains a pressing need for systematic evaluation and multicenter validation of FM outputs within maternal and pediatric populations to ensure they perform reliably in the unique and sensitive contexts of MCH.

### Future Directions

To enable real-world deployment of FMs in MCH, future research must prioritize clinical validation, interpretability, multimodal integration, and domain-specific training. FMs should be evaluated not only retrospectively, but also prospectively in diverse healthcare settings, including rural, low-resource, and multilingual environments, where data are often noisy and incomplete. Integrating structured records, clinical notes, imaging, and time-series data is essential for capturing longitudinal risk trajectories across the maternal-neonatal continuum. Advancing these fields will require interdisciplinary collaboration among clinicians, AI researchers, designers, and ethicists. Co-developing systems that reflect real-world needs and workflows is essential to create tools that are not only technically robust but also socially and clinically acceptable. Ultimately, future FMs should act as intelligent, adaptable allies, which are capable of leveraging heterogeneous data modalities to understand, anticipate, and guide care across all phases of the MCH continuum, thereby advancing truly comprehensive and equitable health outcomes.

## Conclusion

This systematic review offers a comprehensive evaluation of the current landscape of FM applications in MCH, highlighting strong potential of FM to improve MCH care through accurate prediction, clinical evaluation, and information retrieval. They support personalized care, streamline workflows, and enhance decision-making across prenatal, neonatal, and pediatric settings. However, challenges in interpretability, real-world validation, and clinical safety remain. This review highlights both the promise and limitations of FMs in MCH and outlines key priorities for future development, including domain-specific training, multimodal integration, and human oversight. With robust validation, domain-specific training, and interdisciplinary collaboration, FMs can drive a new era of intelligent, equitable, and data-driven maternal and child healthcare.

## Data Availability

The review protocol has been registered on the PROSPERO platform: https://www.crd.york.ac.uk/PROSPERO/view/CRD420251104501 The detailed search strategy and the PRISMA checklist are provided in the Supplementary Material

## Author Contributions

These authors contributed equally: Xinnie Mai and Yunqian Liu

F.X. conceptualized the study and led the work. X.M. and Y.L. conducted literature search. X.M., Y.L., and F.X. screened the titles, abstracts, and full texts. X.M. and Y.L. conducted data synthesis and jointly drafted the initial manuscript, incorporating revisions based on feedback. X.M., Y.L., P.C., J.R., S.Z., M.L., R.W., I.M., M.S., N.A., R.Z., D.S., & F.X. revised the manuscript. F.X. supervised the study. All authors contributed to and approved the final version of the manuscript.

